# Language Model Applications for Early Diagnosis of Childhood Epilepsy

**DOI:** 10.1101/2025.01.31.25321308

**Authors:** Jitse Loyens, Geertruida Slinger, Nynke Doornebal, Kees P.J. Braun, Willem M. Otte, Eric van Diessen

## Abstract

**Objective:** Accurate and timely epilepsy diagnosis is crucial to reduce delayed or unnecessary treatment. While language serves as an indispensable source of information for diagnosing epilepsy, its computational analysis remains relatively unexplored. This study assessed – and compared – the diagnostic value of different language model applications in extracting information and identifying overlooked language patterns from first-visit documentation to improve the early diagnosis of childhood epilepsy.

**Methods:** We analyzed 1,561 patient letters from two independent first seizure clinics. The dataset was divided into training and test sets to evaluate performance and generalizability. We employed two approaches: an established Naïve Bayes model as a natural language processing technique, and a sentence-embedding model based on the Bidirectional Encoder Representations from Transformers (BERT)-architecture. Both models analyzed anamnesis data only. Within the training sets we identified predictive features, consisting of keywords indicative of ‘epilepsy’ or ‘no epilepsy’. Model outputs were compared to the clinician’s final diagnosis (gold standard) after follow-up. We computed accuracy, sensitivity, and specificity for both models.

**Results:** The Naïve Bayes model achieved an accuracy of 0.73 (95% CI: 0.68-0.78), with a sensitivity of 0.79 (95% CI: 0.74-0.85) and a specificity of 0.62 (95% CI: 0.52-0.72). The sentence-embedding model demonstrated comparable performance with an accuracy of 0.74 (95% CI: 0.68-0.79), sensitivity of 0.74 (95% CI: 0.68-0.80), and specificity of 0.73 (95% CI: 0.61-0.84).

**Conclusion:** Both models demonstrated relatively good performance in diagnosing childhood epilepsy solely based on first-visit patient anamnesis text. Notably, the more advanced sentence-embedding model showed no significant improvement over the computationally simpler Naïve Bayes model. This suggests that modeling of anamnesis data does depend on word order for this particular classification task. Further refinement and exploration of language models and computational linguistic approaches are necessary to enhance diagnostic accuracy in clinical practice.

## 1. INTRODUCTION

Epilepsy significantly impacts psychosocial well-being and can adversely affect health-related quality of life.^1,2^ This impact is particularly concerning in children, where recurrent seizures can interfere with normal brain development, potentially leading to cognitive and behavioral impairments.^3–5^ These serious consequences underscore the critical importance of obtaining an early and accurate diagnosis of epilepsy.

Diagnosing epilepsy presents significant challenges due to its polymorphic nature.^6–8^ Research has shown that nearly half of the patients assessed for initial seizures were already experiencing recurrent, undiagnosed seizures at the time of evaluation. While diagnostic time is typically brief for clearly identifiable cases of epilepsy, it can extend beyond a year for complex or ambiguous presentations.^9,10^ This diagnostic uncertainty can have serious consequences: diagnostic delays expose children to ongoing seizures that may impair cognitive development, while false-positive diagnoses can lead to unnecessary administration of antiseizure medications with potential adverse effects.^11–13^

Language plays a fundamental role in epilepsy diagnosis, treatment evaluation, and patient care management. Clinicians rely heavily on patient history and narrative to distill relevant clinical information.^14^ This makes collected text a rich and versatile medium for gaining deep insight into the patient’s condition—an essential component for a comprehensive approach to epilepsy care. Despite advances in ancillary investigations, clinical information from patient records remains indispensable for diagnosing and monitoring epilepsy.^15–17^ However, this wealth of information is often stored in electronic health records in an unstructured manner, limiting its optimal utilization in clinical decision-making.^18^

The emergence of natural language processing (NLP) offers a promising solution for systematically processing this unstructured textual data. NLP, a form of artificial intelligence, specializes in the computational analysis of spoken and written language to identify general patterns and trends and extract relevant information.^19–21^ This involves converting unstructured text into a structured format and applying computational algorithms to analyze these structured features, enabling the retrieval of desired information.

In epilepsy research, there is a growing trend toward NLP applications, including patient identification, risk stratification, and outcome prediction. In clinical settings, NLP can contribute significantly to the early detection and classification of medical conditions, thereby reducing time to diagnosis and treatment.^18^ Recent advances have led to improved NLP models with new generative properties, known as large language models (LLMs).^22–24^ The essence of these models is a transformer architecture with an attention-layer, allowing both an efficient representation and retrieval of relevant information in (textual) data.^24^ Despite the potential of these more advanced language models, their applicability for early diagnosis of epilepsy based on medical documentation remains limitedly explored.^25–27^ This study aims to assess—and compare—the diagnostic value of different NLP approaches using medical letters from first consultations to facilitate the early diagnosis of childhood epilepsy.

## 2. METHODS

### 2.1. Dataset

Our analysis encompassed 1,561 medical patient letters, with 1,250 originating from University Medical Center Utrecht (UMCU) and 311 from Martini Hospital Groningen (MZG). We retrospectively collected data from children (age < 18 years) referred to the First Seizure Clinic (FSC) between 2008 and May 2022. These data were originally collected for previously published studies focusing on prediction model development for childhood epilepsy and the clinical characteristics and diagnoses of children referred to an FSC.^28,29^ Theinstitutional ethics committee of both University Medical Center Utrecht and Martini Hospital approved the use of anonymized retrospective data for research purposes with-out informed consent.

All patient letters were either written and/or supervised by experienced pediatric neurologists. For each patient, we included both the initial diagnosis (established after FSC consultation) and the final diagnosis (reached through consensus among doctors and/or ancillary investigations at the latest follow-up, recorded within a two-year period). Follow-up occurred for children with inconclusive diagnoses at the first consultation and for those initially diagnosed with epilepsy. Children whose epilepsy diagnosis was ruled out were referred back to their referral specialist or general practitioner for follow-up.

Both initial and final diagnoses were categorized into three groups: ‘epilepsy’, ‘no epilepsy’, and ‘unclear’ (**Figure 1**) and served as the model’s outcome. All epilepsy diagnoses were established according to the International League Against Epilepsy definition of epilepsy.^30^ A diagnosis was classified as ‘unclear’ at the initial stage if ancillary investigations were deemed necessary to confirm or reject the epilepsy diagnosis. The final diagnosis was classified as ‘unclear’ if, despite further investigations, uncertainty remained about whether the events were indeed epilepsy-related.^7^

**Figure 1.**
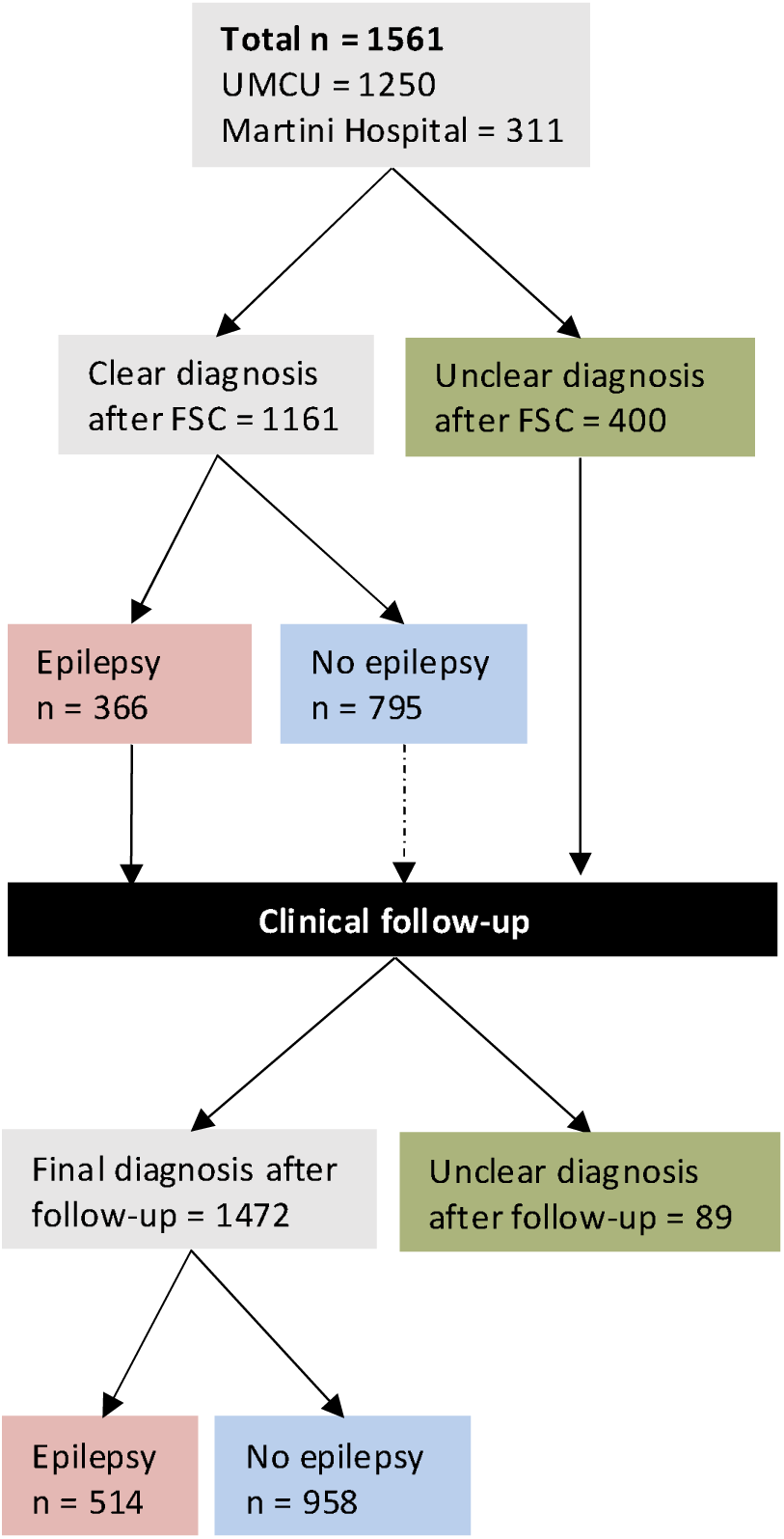
Flowchart illustrating the diagnostic pathway for children referred to the FSC. The flowchart outlines the process from the first FSC consultation to the final diagnosis, including follow-up procedures. The diagnoses are categorized as ‘epilepsy’, ‘no epilepsy’, or ‘unclear’.

### 2.2. Study Design

We conducted a retrospective analysis of the letters to assess the clinical value of language models for early diagnosis of childhood epilepsy. This was achieved through binary text classification, specifically by training classification models based on textual features and predicting the class of new texts within the ‘epilepsy’ and ‘no epilepsy’ patients. To reduce interpretative bias, we exclusively used textual information from patient anamnesis, excluding subjective information from ancillary investigations, conclusions, treatment plans, and clinical considerations.

Our study involved two distinct analyses. **Analysis A**: we combined data from both hospitals and randomly divided it into a training set (80%; 1,173 subjects) and a test set (20%; 293 subjects). To ensure representative distribution of final diagnoses in both sets, we applied stratification based on the final diagnosis groups. **Analysis B**: we created a separate test set comprising all 316 subjects that remained unclear after initial FSC evaluation. This second analysis aimed to determine whether the model could accurately classify initially unclear cases as either having epilepsy or not (**Figure 1**).

The letter corpus exhibited considerable variation in textual length, ranging from 63 to 1,070 words, with a median of 400 words and a mean of 414 words. Four cases (three from the UMC Utrecht; two male subjects) were excluded from the training set due to their succinct nature, consisting of only single sentences in their amnestic report.

### 2.3 Naïve Bayes Model

We used a Naïve Bayes classifier as NLP approach, giving its simplicity and effectiveness in text classification. The essence of the model is the application of Bayes’ theorem, assuming a strong independence between features.^31,32^ Model development contained three phases: data preprocessing, data analysis with feature selection, and classification (**Figure 2**).

**Figure 2.**
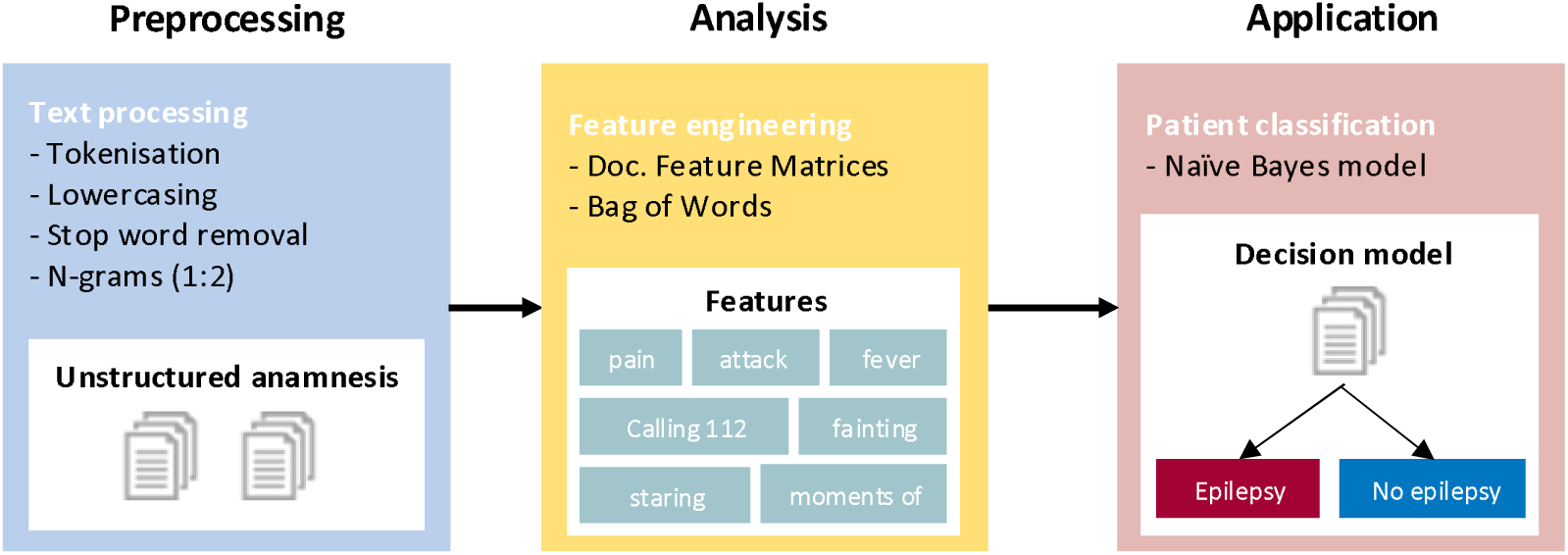
NLP workflow for classifying ‘epilepsy’ or ‘no epilepsy’ diagnosis based on unstructured letters from the first consultations. The process consists of three main stages: preprocessing, analysis, and application.

#### Data preprocessing

Preprocessing encompasses several key steps including corpus creation, tokenization, data cleaning, lowercasing, n-gram generation, and stop word removal. Creating a corpus involves collecting and organizing a substantial amount of textual data in a structured manner to facilitate systematic analysis and processing. Text was then divided into tokens (i.e., words) through tokenization. Undesired characters, such as punctuation marks, symbols, URLs, and separators, were omitted (data cleaning). Lowercasing converted all characters in the text to lowercase letters, ensuring consistency across the tokens. Afterwards, n-grams were generated, with a maximum n-value of 2. N-grams are sequences of consecutive words and will be used as features for the text classification model. It was decided to generate unigrams (single words such as “trekkingen” *(“jerks”)*) and bigrams (pairs of consecutive words such as “geen_trekkingen” *(“no_jerks”)*). The final step involved removing stop words from the generated n-grams. Removing stop words after generating n-grams ensures that some meaningful bigrams are retained, even if they contain stop words (e.g., “geen_koorts” *(“no_fever”)* may be retained while “geen” *(“no”)* and “koorts” *(“fever”)* may individually be stop words). Stop words contain common words including prepositions, personal pronouns, units, and auxiliary verbs that lack informativeness and may interfere with model development. After the preprocessing step, the dataset was split into training and test sets.

#### Data analysis

We created a document-feature matrix (DFM) for the training set to enable structured analysis of text data, representing documents (letters) as rows and features (i.e., all n-grams) as columns. The matrix values represented the frequency of a features in each letter, creating a Bag-of-Words (BoW) model.^33^ In this model the input text is represented as a collection of words, disregarding the order in which they appear. We applied Term Frequency-Inverse Document Frequency (TF-IDF) to weigh features based on their frequency in individual letters. TF-IDF reduces the influence of frequently occurring features while emphasizing more informative ones.

#### Feature selection

Feature selection was achieved through Recursive Feature Elimination (RFE) with 5-fold cross-validation. RFE identified the top 300 features that were most informative for the model’s performance. A selection of 300 features was based on theoretical and practical reasons. Firstly, we wanted to follow the rule of thumb that recommends one feature per ten cases to minimize overfitting and optimize model performance. As the dataset is of medium size, we adjusted this rule to one feature per five cases, resulting in the selection of 300 features. Secondly, the literature supports this selection, as studies frequently use between 200 and 300 features to capture significant patterns while minimizing noise, thereby enhancing the robustness and generalizability of the model. Thirdly, fewer features improve computational efficiency, making the model more practical for implementation. Moreover, fewer features improve the model’s interpretability and transparency, facilitating a better understanding of which variables contribute to its predictions. As a hyperparameter for the Naive Bayes model, the smoothing parameter (α) was added to prevent zero probabilities.

### 2.4. Sentence-embedding Model and Subsequent Classification

This study also employed a classification model to predict epilepsy diagnoses based on patient text records that takes—in contrast to the BoW approach—word sequence into account. First, textual data underwent systematic preprocessing. Initial preprocessing steps included case normalization to lowercase, standardization of special characters to their lexical equivalents, removal of extraneous punctuation marks, and normalization of whitespaces. Next, the processed texts were then embedded using a freely-available multilingual embedding (i.e., the paraphrase-multilingual-mpnet-base-v2 transformer model).^*^ The embedding model implements the Sentence-BERT architecture to generate a contextualized 768-dimensional semantic vector representations for each text, irrespective of its length.^34^ This embedding model was selected for its capacity to preserve both sequential word order information and cross-lingual semantic relationships. Third, the resulting high-dimensional embeddings served as input features for a gradient boosting classifier implemented through the XGBoost framework in R.^35^ The binary classification model employed a linear booster with default hyperparameters, leveraging sequential tree building to iteratively optimize the prediction objective while maintaining computational efficiency.

### 2.5. Performance Evaluation

Performance evaluation utilized confusion matrices to compare actual and predicted classifications through decision statistics from contingency tables. We compared each model’s output to the clinician’s final diagnosis (gold standard). Key performance metrics included: accuracy (i.e., the proportion of correct classifications), sensitivity (true positive rate), and specificity (true negative rate). All evaluation analyses were performed using R software, version 4.4.0.

## 3. RESULTS

### 3.1 Data Characteristics

The median age at the first seizure was 4.5 years (95% CI: 4.0-4.9). The maximum age recorded was 17.8 years, while the minimum age was 1 month. The majority of patients were male, comprising 853 individuals (54.6%). After the first consultation, 366 diagnoses were classified as ‘epilepsy’, 795 as ‘no epilepsy’, and 400 as ‘unclear’. According to the final diagnoses, 514 diagnoses were classified as ‘epilepsy’ (413 from UMCU and 101 from MZG), 958 as ‘no epilepsy’ (767 from UMCU and 191 from MZG), and 89 as ‘unclear’ (70 from UMCU and 19 from MZG). The data characteristics are presented (**Table 1A, Supplementary Materials**).

**Table 1.** Model performance metrics for epilepsy diagnosis: test sets. NB = Naïve Bayes. Between parentheses = 95% Confidence Interval.

| Analysis | Model | Accuracy | Sensitivity | Specificity | PPV | NPV |
| --- | --- | --- | --- | --- | --- | --- |
| A | NB | 0.73<br>(0.68–0.78) | 0.79<br>(0.74–0.85) | 0.62<br>(0.52–0.72) | 0.80<br>(0.74–0.86) | 0.61<br>(0.51–0.70) |
|  | Sentence-embedding | 0.74<br>(0.68–0.79) | 0.74<br>(0.68–0.80) | 0.73<br>(0.61–0.84) | 0.92<br>(0.88–0.96) | 0.39<br>(0.30–0.49) |
| B | NB | 0.74<br>(0.69–0.79) | 0.82<br>(0.77–0.88) | 0.57<br>(0.47–0.66) | 0.79<br>(0.74–0.84) | 0.62<br>(0.52–0.72) |
|  | Sentence-embedding | 0.72<br>(0.67–0.77) | 0.75<br>(0.70–0.80) | 0.57<br>(0.44–0.71) | 0.89<br>(0.85–0.94) | 0.32<br>(0.23–0.41) |

### 3.2. Most Important Features

The *Term Frequency-Inverse Document Frequency* identified several key features most characteristic for the epilepsy classification texts. Notable predictive n-gram feat1ures included: “spray” *(“spray”)*, “kwijlde” *(“drooled”)*, “haar_mond” *(“her_mouth”)*, “insult_doorgemaakt” *(“experienced_insult”)*, “dubbele tong)” *(“slurred_speech”)* and “afgelopen_dagen” *(“last_days”)*. Some features directly reflected clinical observations or descriptions that frequently appear in letters of epilepsy patients, while others, such as “afgelopen_dagen” (“*last_days*”) showed less obvious connection to epilepsy. Lists of the most important features, for the epilepsy as well as the control group, are provided in **Figure 3**.

**Figure 3.**
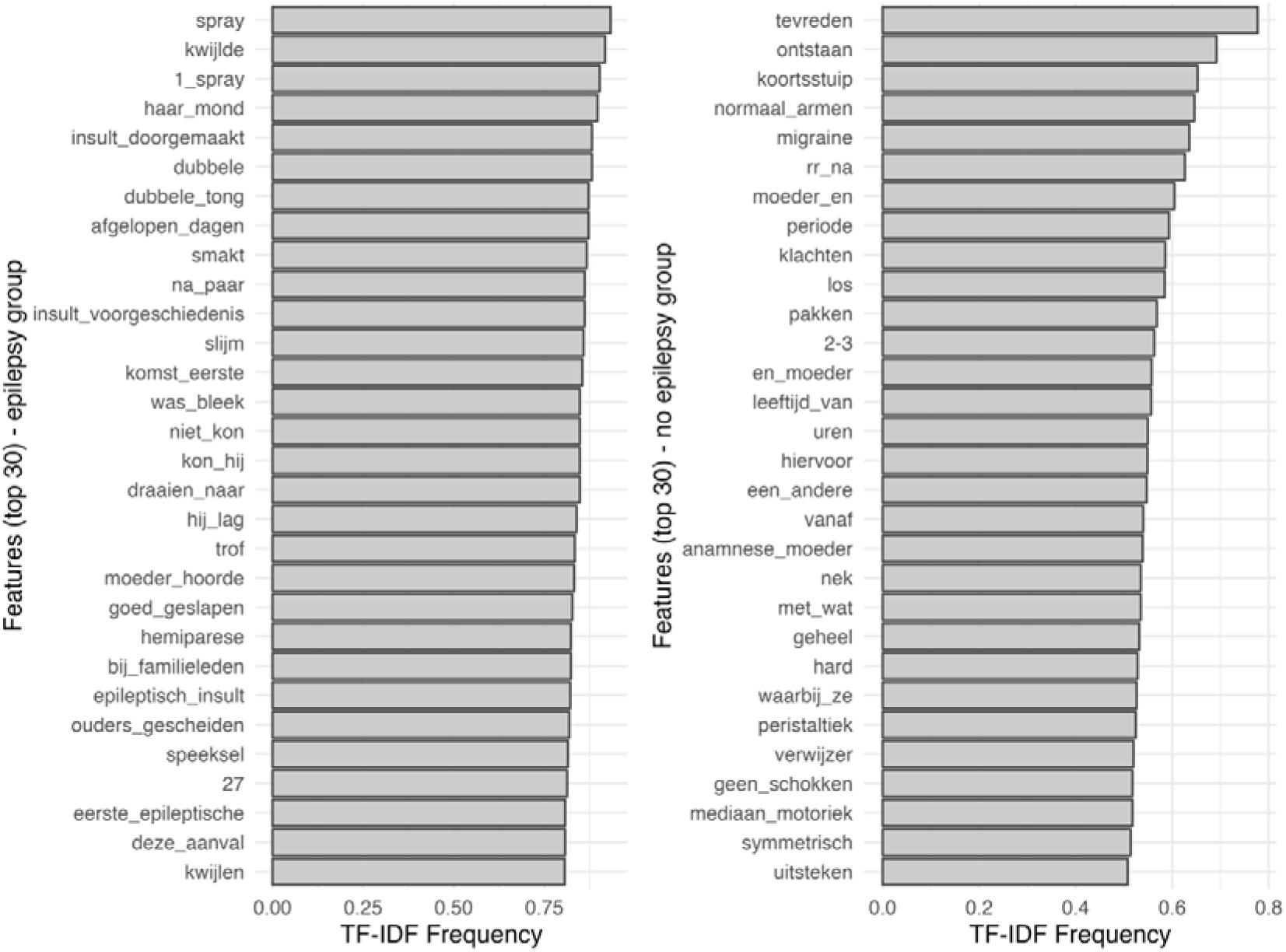
A graphical representation of the most prevalent features for each condition. TF-IDF = Term Frequency-Inverse Document Frequency; to weigh features based on their frequency in individual letters. A complete translation list in English of features is provided (**Table 2A, Supplementary Materials**).

### 3.3. Classification Model Performance

#### Analysis A

The performance of the language models was evaluated on a test set of 293 letters. The Naïve Bayes model correctly identified 62 letters as positive and 153 as negative, resulting in 40 false positives and 30 false negatives, with an overall accuracy of 0.73 (95% CI: 0.68-0.78). The Sentence-embedding model correctly identified 40 letters as positive and 176 as negative, classifying 62 false positives and 15 false negatives, with an overall accuracy of 0.74 (95% CI: 0.68-0.79). An overview including sensitivity, specificity, PPV and NPV is provided (**Table 1**).

#### Analysis B

The performance of the language models was evaluated on a test set of 319 letters with an ‘unclear’ diagnosis. The Naïve Bayes model correctly identified 60 letters as positive and 173 as negative, resulting in 37 false positives and 46 false negatives, with an overall accuracy of 0.74 (95% CI: 0.69–0.79). The Sentence-embedding model correctly identified 31 letters as positive and 196 as negative, classifying 66 false positives and 23 false negatives, with an overall accuracy of 0.72 (95% CI: 0.67–0.77). An overview including sensitivity, specificity, PPV and NPV is provided (**Table 1**).

## 4. DISCUSSION

This study evaluated and compared different language model applications for improving early diagnosis of childhood epilepsy through automated analysis of first-visit documentation. Our findings revealed comparable performance between the simpler Naïve Bayes model and the more advanced Sentence-embedding model, with both achieving moderate to good diagnostic accuracy. Notably, both models demonstrated higher specificity than sensitivity across all analyses, suggesting particular utility in helping clinicians rule out epilepsy diagnoses and identify cases requiring additional investigation. Previous research has established the value of NLP in various aspects of epilepsy care, including patient identification,^36–38^ information retrieval,^39–41^ and coping strategies.^42,43^ Recent studies have begun exploring language applications in early clinical phenotyping and genetic epilepsies.^44,45^ However, our study uniquely addresses the specific challenges of early childhood epilepsy diagnosis, where textual analysis holds particular promise given the heterogeneous presentation of symptoms.

The performance metrics should be considered in context of the model performance: our models relied solely on patient narratives, deliberately excluding information from EEG reports, clinical evaluations, and medical conclusions. From this perspective, the application of language models could even be used in the early phase of clinical evaluation of child suspected of epilepsy. Interestingly, the transformer-based Sentence-embedding model – which takes word order into account – demonstrates no significant improvement over the Naïve Bayes model. The Naïve Bayes model is regarded as a robust classification model, even when working with limited data and feature sets.^31^ Transformer-based language models perhaps require longer text sequences to effectively recognize desired patterns, particularly in cases with less variation in language utilization. With limited text input, simpler models can offer greater practical value in practice, where speed, simplicity and apprehensibility often take precedence in the implementation within clinical workflows. The achieved accuracy levels suggest potential value for early-stage screening and decision support.

Interestingly, our study revealed that both obvious as less obvious words (or combinations) are of additional value for correct classification. The use of epilepsy-related terminology could reflect the physician’s (implicit) evaluation of the clinical case during consultation. Unrelated word (combinations) with no obvious relation to epilepsy that were classified as relevant features for model develop may represent either underlying linguistic patterns common in epilepsy-related letters, or potential limitations in the model’s feature selection process. Previous efforts in the field have revealed similar insights into the non-semantic evaluation of patient history, and showed that hesitations and formulation efforts might be of additional value when diagnosing epilepsy.^46,47^ Future research efforts should therefor a comparison of different language model approaches to further elucidate the true value of these implicit language information for diagnosing epilepsy.

This study benefits from a substantial and diverse dataset collected from two hospitals, enhancing the robustness and generalizability of the model’s results. The retrospective nature and moderate size of our dataset are, however, potential contributors to the limited sensitivity and PPV of both models, thereby increasing the chance of missed epilepsy cases and false positives. Furthermore, performance was significantly higher on the training sets (not reported) compared to the test sets, indicating potential overfitting. Overfitting occurs when the model learns the textual details and noise in the training data, which impairs its generalizability to new data. This can result from excessive noise, an excessive number of features, irrelevant features, or insufficient training data. Equally important to consider is the imbalanced dataset we used (uneven class distribution), predominantly consisting of letters from children with a ’no epilepsy’ diagnosis. Imbalanced data can hinder a model’s ability to learn the minority class, as (language) models often exhibit a preference for the majority class.^48^

From a model perspective, few limitations should be mentioned. A Naïve Bayes model is a relatively limited in its capacity to learn complex (textual) relations.^31^ The model does not adequately account for word order or combinations of words, potentially resulting in misinterpretations of negations and the overall meaning within clinical text. Confounding factors like typographical errors, abbreviations, double negations, and letters written by multiple authors can adversely affect the classification process. Additionally, RFE was applied to a subset of the top 8000 features (i.e., 300) due to computational constraints, possibly excluding relevant features. A general limitation of feature selection is the possible omission of rare but significant features, particularly in the context of rare diseases or syndromes. Technically, more word-order-oriented models could (partially) overcome the aforementioned model limitations due to their transformer-architecture in which meaningful textual relations are represented internally. Mechanism that drives these models to achieve such model properties remain difficult to grasp, prohibiting a better of understanding of these models.^49,50^

Future research should incorporate a prospective design to explore the clinical applicability. Prospective studies enhance variable control, minimize data noise, and allow real-time language capture, thereby reducing biases and missing data. This would also allow to capture a recorded – instead of a written – patient history that would inevitably lead to new potential hidden language domain sources (e.g., phonology, prosody, syntax use) to improve epilepsy diagnosis.^46,47^ This could be particularly beneficial for LLMs as these models excel in retrieving ‘hidden’ textual association that might be use for classification. Enhancing algorithms, refining feature selection, and utilizing larger, more diverse datasets are essential to improve diagnostic accuracy. Apart from methodological improvements, integration of language-based classification models with existing clinical diagnostic tools in epilepsy care would be a next step to explore its actual clinical value.^28,51,52^

## 5. CONCLUSIONS

Our study demonstrates that both simple and complex language models can achieve meaningful performance in supporting early childhood epilepsy diagnosis, even when limited to first-visit documentation. The comparable performance between Naïve Bayes and more sophisticated transformer-based language model suggests that simpler, more interpretable models may be preferable for initial clinical applications as long as the input data is limited in size and complexity. While further refinement is needed, these findings support the potential value of computational linguistic approaches in improving early epilepsy diagnosis and patient care. The higher sensitivity demonstrated by both models suggests particular utility in helping clinicians identify cases that do not require additional investigation, potentially streamlining the diagnostic process and reducing unnecessary testing. As these methods continue to evolve, their integration into clinical practice could provide valuable decision support for clinicians while maintaining the essential role of clinical expertise in final diagnostic decisions.

## Data Availability

All data produced in the present study are available upon reasonable request to the authors

## Appendix

**Table A1.**
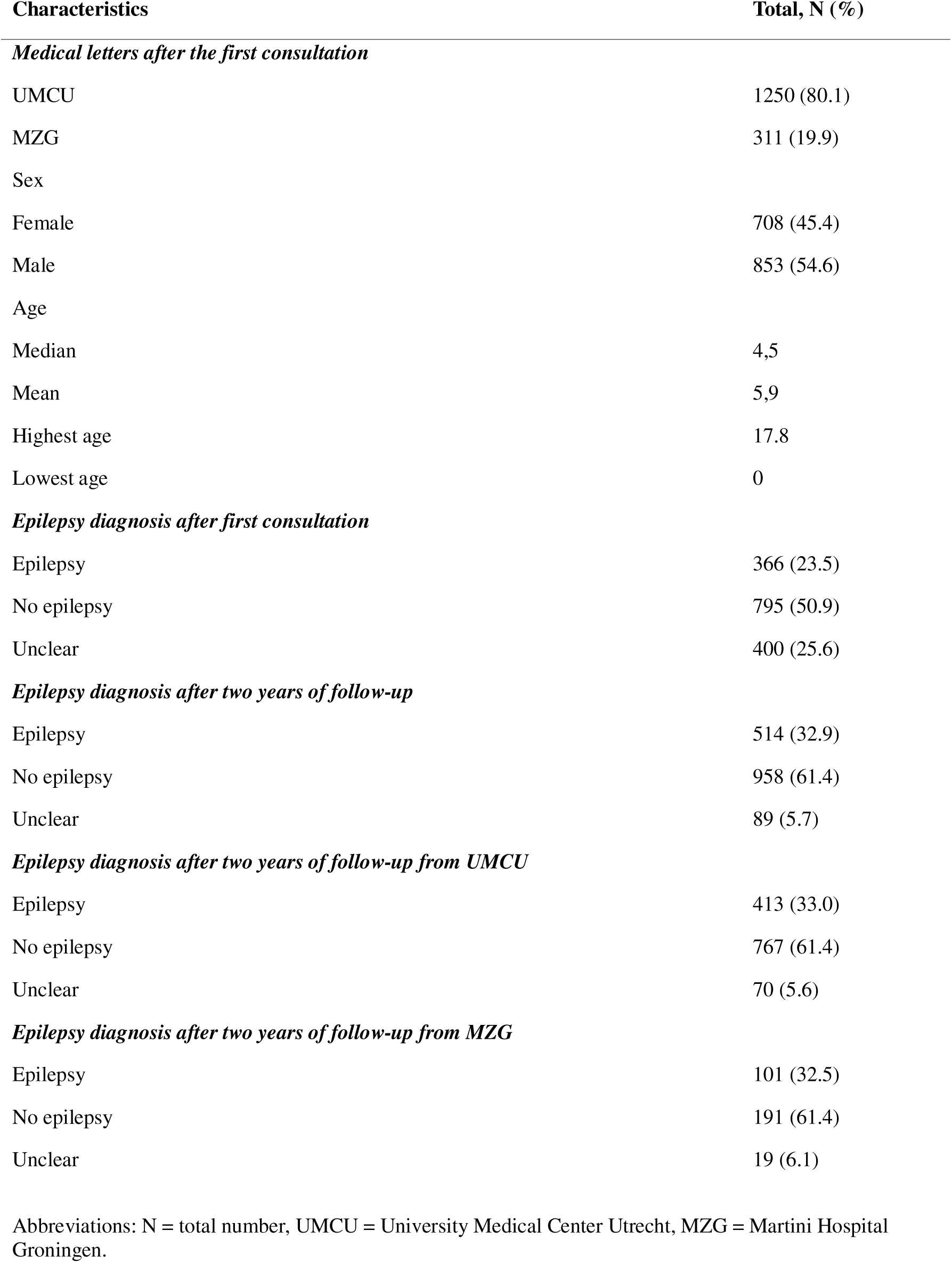
Baseline characteristics of the data.

**Table A2.**
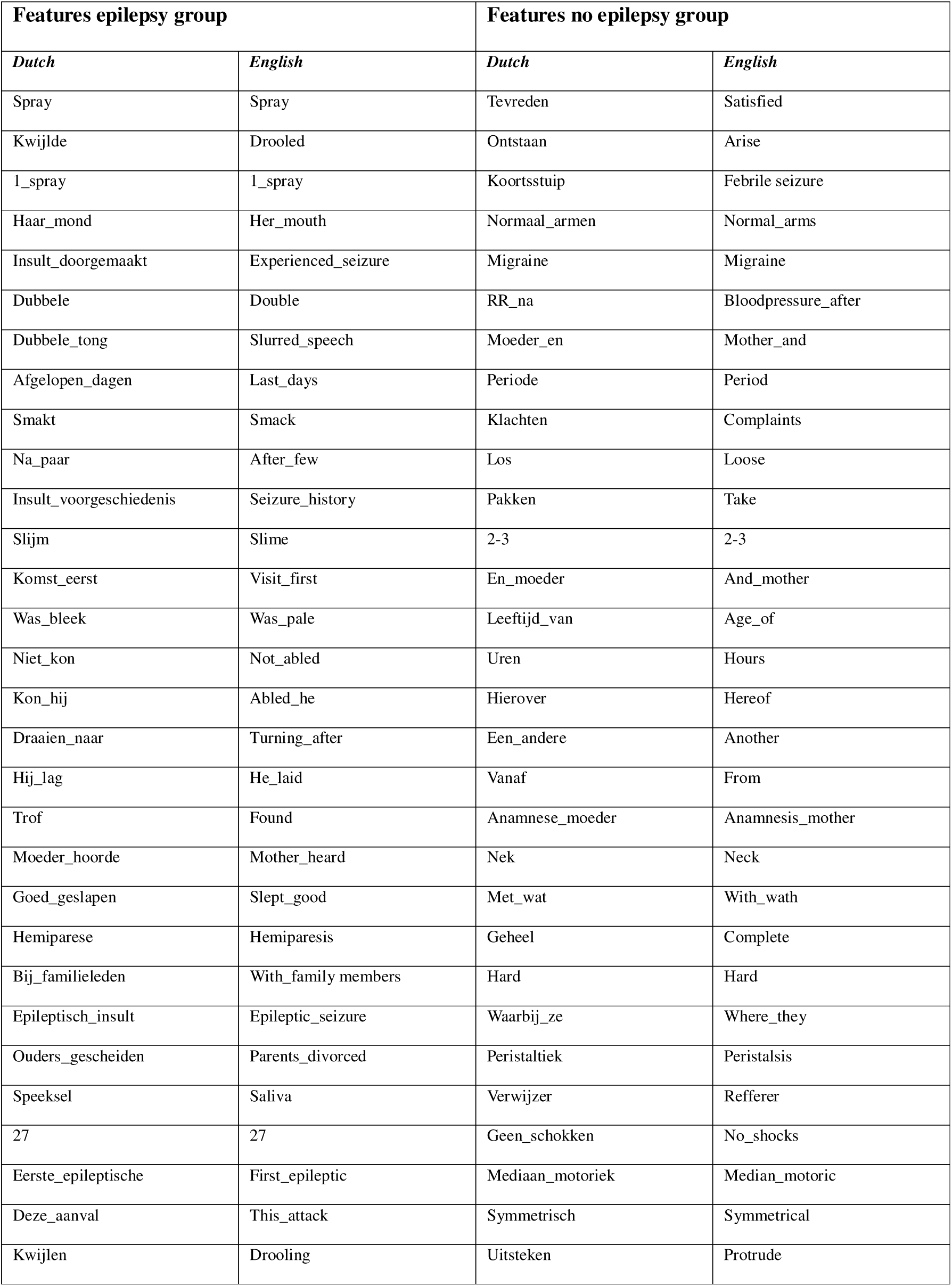
A complete translation list of most prevalent features (Dutch-English)

## Footnotes

* https://huggingface.co/sentence-transformers/paraphrase-multilingual-mpnet-base-v2

